# Evaluation of liver function tests and C-reactive protein in COVID-19 (SARS Cov-2) positive patients diagnosed by Real-time PCR

**DOI:** 10.1101/2021.09.29.21264304

**Authors:** Fatima Khurshid, Sajjad Iqbal, Madiha Mumtaz

**Author notes:** CORRESPONDING AUTHOR EMAIL ADDRESS:* Fatima Khurshid. DECLARATION OF AUTHOR’S COMPETING INTERESTS:* The authors declare that they have no known competing financial interests or personal relationships that could have appeared to influence the work reported in this paper.

## Abstract

**BACKGROUND AND AIMS:** The SARS-CoV-2 pandemic, has caused an unconventional social and economic impact globally. To date, there was limited data regarding the effect of COVID-19 infection on the trend of RT-PCR Ct value, risk factors for disease, effect on liver enzymes, etc. This study aimed to assess the frequency of COVID-19 infection in different age groups and genders. Association of cycle threshold (Ct) values with disease severity and to describe the effect of COVID-19 infection on LFT, Deritis ratio, and CRP. That can be used as indicators for COVID-19 infection diagnosis, the guidance for treatment decisions, and prognosis in infected individuals.

**METHODS:** This was a cross-sectional study conducted in the Molecular Biology and Chemical Pathology sections of the Pathology Department, Shalamar Teaching Hospital Lahore from November 2020 to March 2021.

**Results:** Males 51% were more likely to be infected by SARS-CoV-2. Most of the infected individuals 36.5% were in the age group 20-40. Age and underlying comorbidities are important factors that play a significant role in COVID-19 severity. The uppermost number of the patients had symptoms of fever 78.3%, cough 50.4% and myalgias 50.1% RT-PCR low Ct value could be an important indicator related with the disease severity and mortality risk p value < 0.001 and 0.003 respectively. Bilirubin indirect, ALT, AST, and CRP were significantly associated with disease severity. Deritis ratio and CRP was found to be significantly associated with the risk of mortality.

**CONCLUSIONS:** Real-Time PCR results along with Ct values for SARS-CoV-2 may have benefit for clinicians in patient management decisions. Several risk factors e.g., age and comorbidities for developing severe disease and mortality risk have been identified. These biochemical laboratory parameters ALT, AST, Deritis ratio and CRP can be used as predictive biomarkers for progression towards severe disease and risk of mortality.

## INTRODUCTION

Before 2019 ends, an obscure infection that causes pneumonia like symptoms and lung fibrosis arose. The outbreak was originated in Wuhan City, Hubei province of China. This pneumonia of unknown etiology was reported to World Health Organization (WHO) on 31^st^ December 2019.^(1, 2)^ Initially WHO named it as 2019-novel coronavirus (2019-nCoV) on January 12, 2020 after that on the basis of phylogenetic analysis International Committee on Taxonomy of Viruses officially named it as severe acute respiratory syndrome 2 (SARS-CoV-2) on 11 February 2020.^(3, 4)^ It continues to spread around the globe, and within 5 months, more than 2.6 million people are infected with this virus.^(5)^ This SARS-CoV-2 was declared as a pandemic by WHO on 11 March 2020. It was the fifth pandemic after the flu in 1918.^(4, 6)^

The coronavirus has been previously known for causing Severe Acute Respiratory Syndrome (SARS-CoV) and Middle East Respiratory Syndrome (MERS-CoV) outbreaks respectively.^(7)^ The novel coronavirus that is emerged recently, is responsible for coronavirus disease 2019 (COVID-19), and named Severe Acute Respiratory Syndrome Coronavirus-2 (SARS-CoV-2) is thought to be more infectious as compared to SARS and MERS-CoV.^(7)^

Coronavirus disease (COVID-19) is a leading public health concern these days.^(8, 9)^ Pakistan, alongside many other countries is also confronting the historic public health emergency these days.^(7)^ The classical symptoms of COVID-19 are fever, pneumonia, dry cough, and myalgia, and even though can cause alveolar damage which can lead to progressive respiratory failure.^(10)^ About 81% of patients of SARS-CoV-2 had asymptomatic or mild disease, 14% progress to severe disease can present with pneumonia or respiratory distress and need medical care, and just 5% of hospitalized patients need intensive care units.^(7)^

The estimated mean incubation period for SARS-CoV-2 is 4 to 6 days, as compared to SARS-CoV in which the mean incubation period was 4.4 days and 5.5 days in MERS-CoV respectively. It is also observed that out of 10,000 infected individuals 64 may develop symptoms after the quarantine period of 14-day. Present studies suggest that older individuals and those with weak immune systems from underlying comorbidities are more prone to develop severe forms of COVID-19.^(11)^

Person-to-person contact and respiratory droplets are the primary sources of SARS-CoV-2 transmission.^(10)^ The inhaled virus binds the epithelial cells present in the nasal cavity and starts replication. The primary receptor for 2019-nCoV is angiotensin-converting enzyme 2 (ACE2) that is a metalloprotease and expressed in the lung, cardiac, liver, intestine, vascular endothelium, and kidney cells.^(8, 12)^

Rapid detection of SARS-CoV and SARS-CoV-2 is done through nucleic acid amplification test (NAAT) by real-time reverse-transcription polymerase chain reaction (RT-PCR) or Enzyme immunoassay (EIA) for antigen detection. Because Serum IgA, IgM, and IgG and will appear simultaneously, between days 5 and 17 afterward the onset of symptoms but the gold standard for diagnosis is RT-PCR.^(12, 13)^ Throat washings and nasopharyngeal swabs preserved in viral transport medium (VTM) remain the key diagnostic samples for PCR diagnosis.^(12, 14)^

In RT-PCR the cycle threshold (Ct) values denote the amplification cycles number, which is necessary for the target gene to surpass a threshold level. It is the indirect method of quantifying the viral RNA copies number in the sample or in other words a semi-quantitative measure of the viral load so Ct value had an inverse relationship with the viral load. Nevertheless, the Ct value is only a proxy for viral load because several factors e.g. the assay itself or the factors in the sample matrix can affect the amplification efficiency.^(15)^

The genome of SARS-CoV-2 codes for four main structural proteins: spike (S), membrane (M), envelope (E), nucleocapsid (N) of which the spike (S) protein plays a major part in receptor recognition and the cell membrane fusion. The S protein typically present in a prefusion, metastable form when the virus connects with the cells of host extensive structural change occurs in the S protein that allows the virus to fuse with the cell membrane of the host.^(16, 17)^ Interestingly, patients diagnosed with COVID-19 infection also had variable grades of hepatic damage/dysfunction besides the acute respiratory symptoms.^(18)^

Involvement of the liver in COVID-19 could be due to an unrestrained immune response, direct cytopathic effect of the virus, drug-induced hepatic injury, or sepsis. ACE2 receptors are generously distributed in type 2 alveolar cells and the suggested mechanism of virus entrance is also through ACE2 receptors of the host. Interestingly, ACE2 receptors are also present in the vascular endothelium, enteric tract, and liver cholangiocyte. Most of the patients develop elevated transaminases regardless of the presence of ACE2 receptors in cholangiocytes.^(19)^

The C-Reactive Protein (CRP) induced by interleukin-6 (IL-6) in the liver is a non-specific acute-phase protein, and is a sensitive biomarker of inflammation, tissue damage and infection. The expression level of CRP is generally low but increases spontaneously and significantly during an acute inflammatory response. Elevated CRP in isolation or conjunction with other parameters may indicate a viral or non-viral infection. The inflammatory response plays an important role in COVID-19 infection and inflammatory cytokine storm also increases the severity of infection. ^(14)^

This is the era of life-threatening viral respiratory infections where human beings are at the leading edge in dealing with such viruses as SARS, MERS, avian influenza H5N1, and H7N9.^(20)^ Recently emerged SARS-CoV 2 has become a public health emergency now a days. Possible signs and symptoms, clinical outcomes, and variations in biochemical markers in our population, that can be used as potential indicators for the disease and those may help the clinicians in diagnosis and direction for the appropriate treatment.

## METHODS

This was a cross-sectional study among patients of Shalamar Teaching Hospital, Lahore. From November 2020 to March 2021 total 345 tested positive for COVID-19 infection by RT-PCR were included in the study. All patients provided informed consent. It was not appropriate or possible to involve patients and the public in the design, conduct, or reporting of our research.

### Liver function test (LFT) and C-Reactive Protein (CRP)

Blood samples of all patients were taken from the venous blood by phlebotomists. Centrifugation was performed at 5000 rpm for 10 minutes to obtain serum. LFT, CRP was performed by photometric and particle enhanced immunoturbidimetric assay respectively. LFT and CRP abnormalities were defined as the elevation or reduction from the mentioned ranges: bilirubin direct (0.10 - 0.40 mg/dl), bilirubin indirect (0.10 – 0.70 mg/dl), alkaline phosphatase (ALP) (40.00 – 135.00 U/L), alanine aminotransferase (ALT) (5.00 – 40.00 U/L) aspartate aminotransferase (AST) (5.00 – 40.00 U/L), albumin (3.50 – 5.00 g/dl), proteins total (6.30 – 8.30 g/dl), gamma G.T (GGT) (5.00 – 50.00 U/L), deritis ratio was calculated AST/ALT (1.00), CRP > 5.00mg/dl.

### Statistical analysis

Categorical variables were described as frequency and percentages, and continuous variables as mean and SD. Comparison of categorical variables was done using the Chi-square or the Fisher exact test if the cell counts were small. One way ANOVA was used to explore the association between liver test abnormalities and the severity of the disease. A p value of less than 0.05 was considered statistically significant. All statistical analyses were performed using SPSS version 22.0 software.

### Ethical Approval

The study was approved by the Institutional Ethical Review Board, Shalamar Medical and dental college (SMDC-IRB/AL/60/2020).

## RESULTS

A total of 345 COVID-19 patients confirmed by RT-PCR were included in this study. Mean Ct value RT-PCR was 15.2±6.60. Male’s patients were 51% (N=176) while 49.0% (N=169) were females. The mean age was 47.5±17.3 years. Most of the infected individuals 36.5% were in the age group 20-40 (Figure 1). The highest number of the patients had symptoms of fever 78.3%, cough 50.4% and myalgias 50.1% (Figure 2). All the patients were classified into mild, moderate, and severe categories on the basis of symptoms as per WHO interim guidance 27 may 2020. The highest percentage of patients 47.02% were in the mild category whereas, 33.30% moderate, 10.70% asymptomatic, 7.70% were severe. Of 345 patients 117 had underlying comorbidities, most of the patients were diabatic 15.4%, hypertensive 9.6%, 5.80% cardiac patients, 2.00% asthma, 1.60% have chronic kidney disease and 0.30% had tumors. Table 1 shows that age and underlying comorbidities (diabetes, hypertension, heart and kidney disease, tumors) are important factors that play a significant role in COVID-19 severity. Besides that, RT-PCR Ct value also has a significant association with disease severity p value <0.001. However, gender takes no role in COVID-19 severity. As of 345 patients, 19 (5.5%) died and 326 (94.5%) recovered. Of patients who died 5.2% had underlying comorbidities, 3.76% were females and 1.73% were males. There was a significant association of age, RT-PCR Ct, underlying comorbidities p value 0.001, 0.003, and < 0.001 with the risk of mortality. However, we found no significant role of gender in disease mortality p value=0.065.

**Figure 1.**
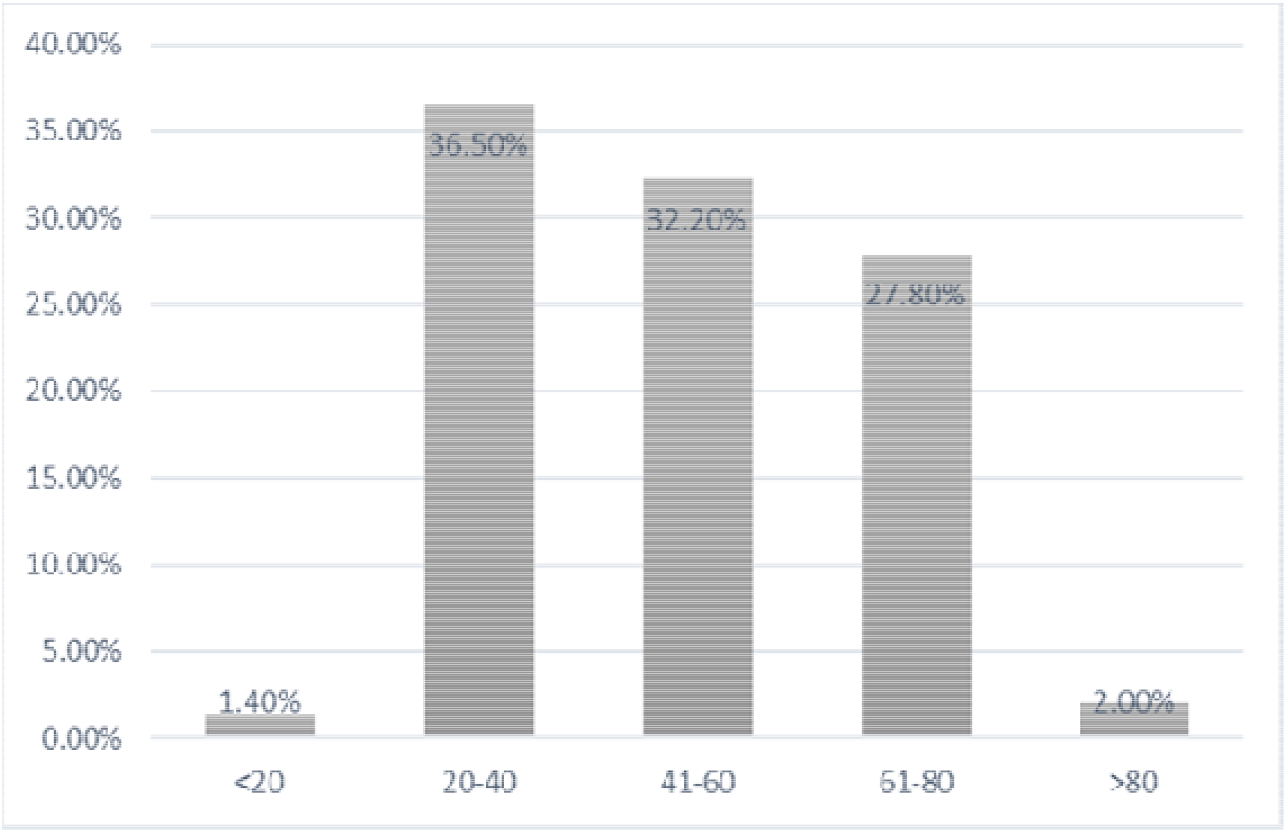
Age Distribution

**Figure 2.**
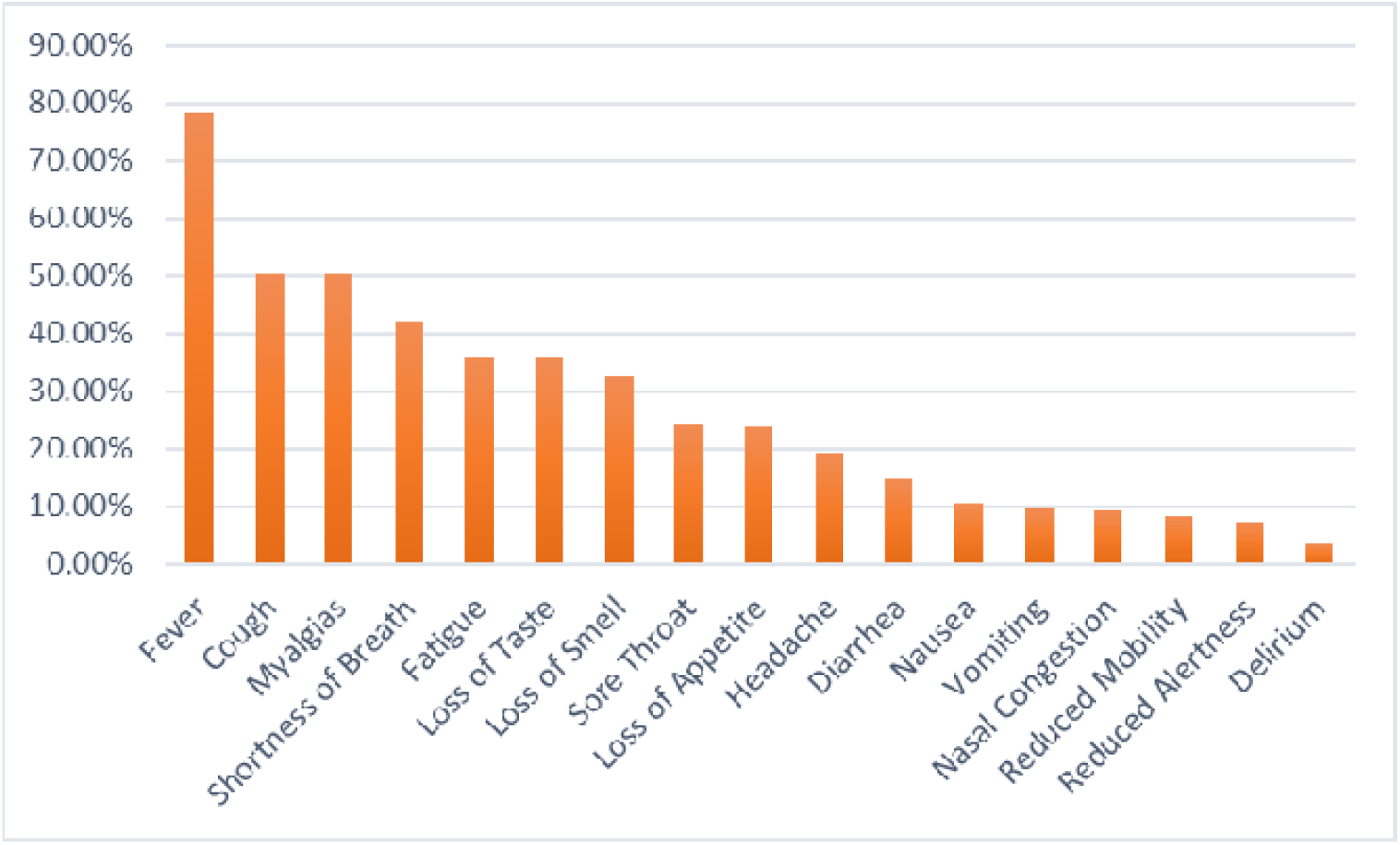
Signs and Symptoms of Disease

**Table 1.**
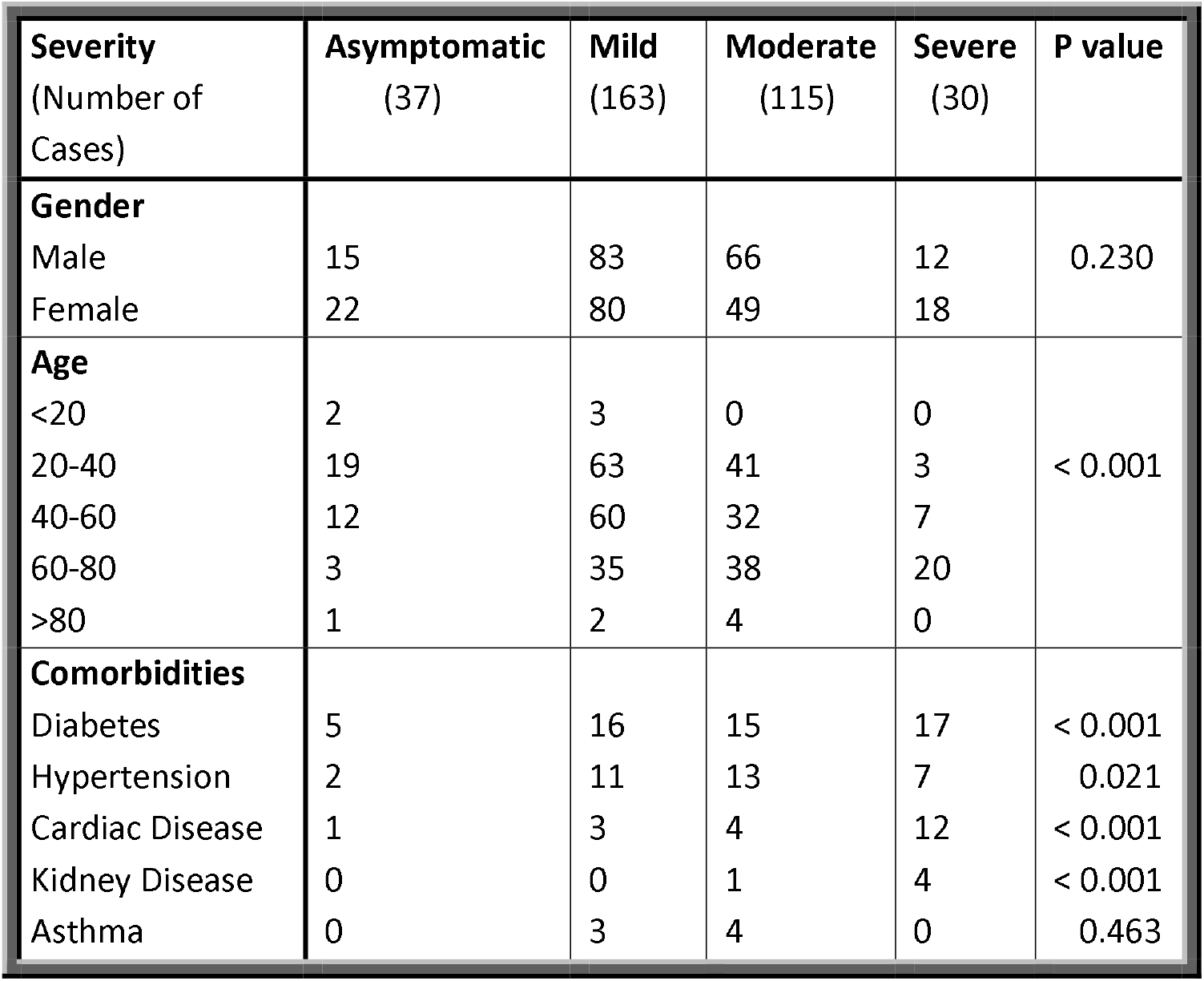
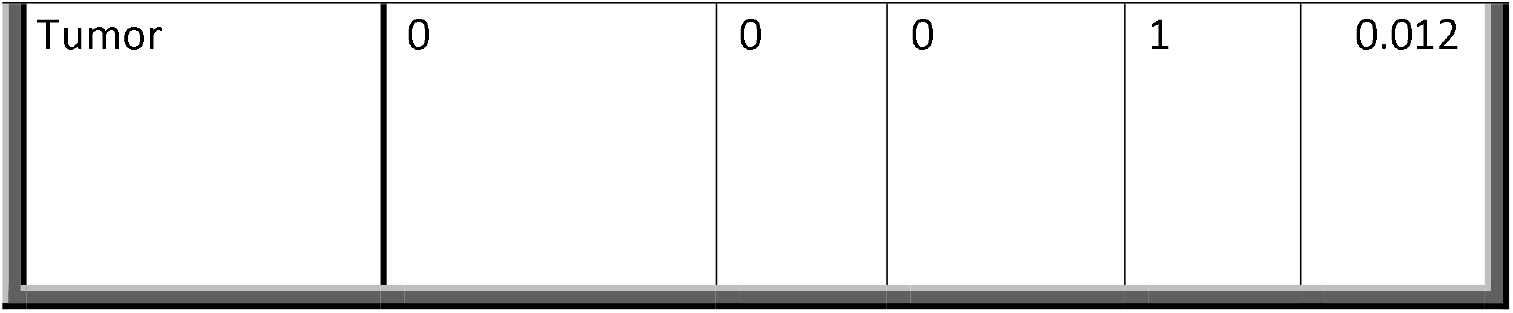
Association of gender, age, comorbidities with COVID-19 severity

| **Severity**<br>(Number of Cases) | **Asymptomatic**<br>(37) | **Mild**<br>(163) | **Moderate**<br>(115) | **Severe**<br>(30) | **P value** |
| --- | --- | --- | --- | --- | --- |
| **Gender** |  |  |  |  |  |
| Male | 15 | 83 | 66 | 12 | 0.230 |
| Female | 22 | 80 | 49 | 18 |  |
| **Age** |  |  |  |  |  |
| <20 | 2 | 3 | 0 | 0 | < 0.001 |
| 20-40 | 19 | 63 | 41 | 3 |  |
| 40-60 | 12 | 60 | 32 | 7 |  |
| 60-80 | 3 | 35 | 38 | 20 |  |
| >80 | 1 | 2 | 4 | 0 |  |
| **Comorbidities** |  |  |  |  |  |
| Diabetes | 5 | 16 | 15 | 17 | < 0.001 |
| Hypertension | 2 | 11 | 13 | 7 | 0.021 |
| Cardiac Disease | 1 | 3 | 4 | 12 | < 0.001 |
| Kidney Disease | 0 | 0 | 1 | 4 | < 0.001 |
| Asthma | 0 | 3 | 4 | 0 | 0.463 |
| Tumor | 0 | 0 | 0 | 1 | 0.012 |

Out of 345 COVID-19 cases confirmed by RT-PCR; ALT, AST and Gamma G.T was deranged in 31.10%, 45.60%, 40.80% patients respectively. However, Albumin 35.90% and proteins total 2.90% found low in most of the cases. Other parameters e.g., bilirubin total, bilirubin direct, bilirubin indirect were deranged in the small number of patients (Figure 3).

**Figure 3.**
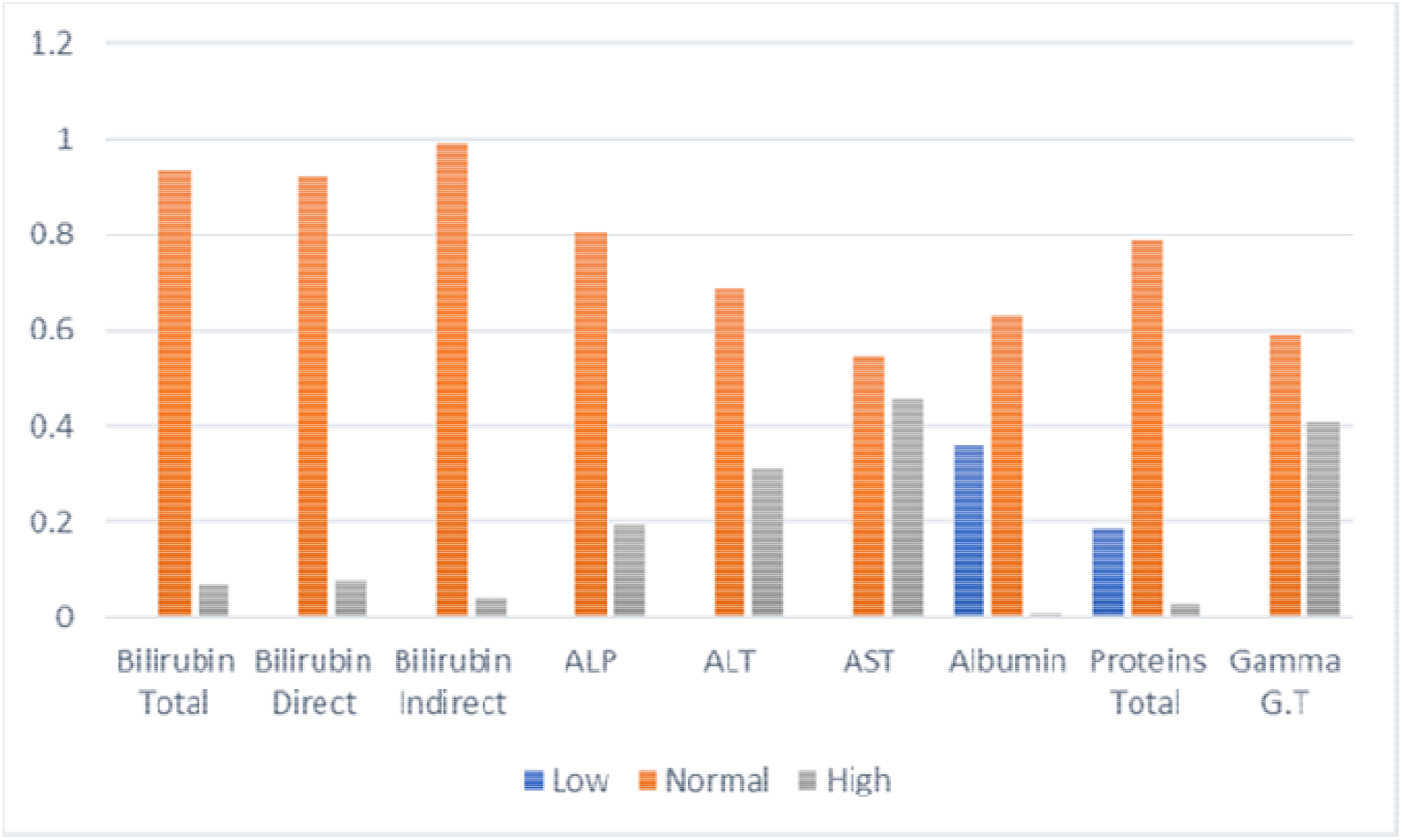
Graphical representation of LFTs parameters in COVID-19 patients

Deritis Ratio It is defined as the activity of AST with ALT (AST/ALT). Only 24.30% of patients had normal deritis ratio <1.00 whereas, it was found raised in 75.70% of patients.

CRP were raised in 78.90% and 21.105% had CRP in normal range.

Table 2 demonstrates the differences for bilirubin (total, direct, indirect), ALP, ALT, AST, albumin, proteins total, GGT, and CRP between asymptomatic, mild, moderate, and severe disease. One way ANOVA result shows that bilirubin indirect, ALT, AST CRP were significantly associated with disease severity.

**Table 2.** Results of ANOVA along with Mean ± SD of Liver Function Test and CRP

| Liver Function Test | Asymptomatic | Mild | Moderate | Severe | P value |
| --- | --- | --- | --- | --- | --- |
| Bilirubin Total (mg/dl) | 0.5182 ± 0.2316 | 0.6842 ± 1.00553 | 0.5028 ± 0.35736 | 1.2444 ± 1.88905 | 0.084 |
| Bilirubin Direct (mg/dl) | 0.2455 ± 0.10357 | 0.2947 ± 0.44048 | 0.2694 ± 0.25837 | 0.7778±1.47471 | 0.053 |
| Bilirubin Indirect (mg/dl) | 0.2727 ± 0.15551 | 0.2474 ± 0.11327 | 0.2333 ± 0.1069 | 0.4667 ± 0.66686 | 0.042 |
| Alkaline Phosphatase (U/L) ALP | 137.2727 ± 82.91573 | 94.6316 ± 51.86803 | 111.1944 ± 54.21324 | 120.8333 ± 103.009 | 0.244 |
| Alanine Amino Transferase (U/L) ALT | 21.6364 ± 9.8313 | 26.3684 ± 16.60742 | 77.3333±142.938 | 70.2778 ± 32.05841 | 0.041 |
| Aspartate Amino Transferase (U/L) AST | 21 ± 5.72713 | 31.9474 ± 13.13742 | 106.0556 ± 201.9209 | 94.8333 ± 69.16753 | 0.033 |
| Albumin g/dl | 4.1182 ± 0.40204 | 4.7395 ± 5.39868 | 3.5639 ± 0.60245 | 3.5667 ± 0.53358 | 0.431 |
| Proteins Total g/dl | 7.0182 ± 1.73195 | 6.9079 ± 0.63219 | 6.8806 ± 0.76525 | 6.8611 ± 0.77698 | 0.968 |
| Gamma G.T (U/L) | 111.1818 ± 164.7136 | 63.1316 ± 64.25505 | 84.6389 ± 73.76223 | 54.1111 ± 89.2148 | 0.26 |
| C- Reactive Protein (mg/dl) CRP | 2.2400 ± 0.61074 | 68.0456 ± 82.06001 | 90.2222 ± 101.32258 | 143.6524 ± 100.00946 | 0.002 |

Deritis ratio and CRP were also found to be significantly associated with the risk of mortality p value 0.018 and 0.030 respectively.

## DISCUSSION

This study summarizes the RT-PCR Ct/ the viral load of the disease, clinical history, disease severity and other biochemical parameters such as LFTs, Deritis ratio, and CRP in COVID-19 In the present study 51% males and 49 % females affected with the disease the same results was observed in Jordan et al., 2020 study. According his study (53.4%) males infected with SARS-CoV-2. Males were more susceptible to SARS-CoV infection as compared to females which could be due to their exposure, higher smoking habits and subsequent comorbidities.^(21-23)^ Mean age was 47.5 years and 36.5% was in age group 20-40 in our study whereas 49.0-48.0 years in previous studies.^(23, 24)^ CDC Director Dr. Rochelle Walensky stated that “hospitals are seeing more and more younger adults” scientists said that new variants of COVID-19 are more transmittable^(25)^ or it could due more exposure of individuals this age with others. Patients with severe disease shows high viral load, low RT-PCR Ct value. Currently, available confirmatory test for COVID-19 is RT-PCR also considered as gold standard to rule out infection in COVID-19 patients. RT-PCR is very specific, sensitive and rapid technique to detect SARS-CoV-2 RNA.^(26)^ The Ct value of RT-PCR denotes the amplification cycles number that are required by the target gene to surpass the threshold level. Previous studies also reported that RT-PCR low Ct value in SARS-CoVcould be an important factor related with the disease severity and mortality risk^(15)^ as in our study with p value < 0.001 and 0.003 respectively.

COVID-19 infection is very heterogeneous it can vary from asymptomatic to mild, moderate, or severe infection. Other than that, the infection can progress either to way worsening of disease and death or improvement and recovery of the patients. In our study found a significant association between high Ct value and severity of disease, mortality risk. Most of the individuals affected by COVID-19 only had mild symptoms as in our study 47.20% were mild cases besides that some patients remain asymptomatic 10.70% during infection. Data from China, also suggested that 81% infected Covid-19 patients had mild or moderate disease, and 14% progress to severe disease.^(27)^ The reason behind this could be their immune status or age as data of case fatality rate (CFR) from Italy which was the first country affected with the pandemic afterwards China.^(28)^ In Pakistan the proportion of people with age 65 years and above was 4.32% as reported^(29)^ but the higher CFR then China (5.5 vs 2.3 %) could be due the availability of advance medical facilities. Global reports shows that almost 60% of deaths were in males^(30)^ that was higher than our study findings where death rate in males were 31.5%.

High risk group of COVID-19 infection and factors that are associated with severe disease includes individuals with age >70 and those with underlying comorbidities e.g. diabetes, hypertension, cardiac, liver, respiratory, kidney diseases or chronic lung disease and tumors.^(21, 31)^ As previously reported by Li et al and CDC that people with hypertension are more at risk for severe COVID-19 (OR: 2.01; P = .003) as in our study we also found diabetes (15.40%), hypertension (9.60%), cardiac disease (5.00%), asthma (2.00%), kidney disease (1.60%) and tumor (0.30%) of patients.^(23, 31)^ Identification of the risk factors that promote the progression towards severe COVID □ 19 have great significance for clinicians and for public health policies to deal with the disease.

The most common clinical symptoms of disease observed in this study were fever 78.3%, cough 50.4%, myalgias 50.1%, and shortness of breath in 42.00% while less common symptoms were headache 19.1%, diarrhea 14.8%. Our study shows similar results reported in previous studies by Huang et al and co-workers also reported in his study the most common symptoms were fever 98%, cough 76%, myalgias 44%, shortness of breath 55% and headache 8%, diarrhea 3% observed as least common symptoms.^(23, 24)^ Elevated levels of ALT (39.4%), and AST (28.1%), also abnormal levels of total bilirubin, lower albumin (2.6 vs. 3.4 g/dl, p<0.01) were reported previously.^(31-34)^ However, we found significantly high levels of bilirubin indirect (P=0.042) transaminases ALT 31.10% (P=0.041) and AST45.60 % (P=0.033) in our findings. Albumin was also found to be low in 35.90% but the association was no significant (P=0.431). The reason for liver damage is still unclear, but some studies show that it could be due to collateral damage due to cytotoxic response during infection as ACE2 receptor the suggested mechanism of virus entrance also present on cholangiocytes, or due to the toxicity from frequently used drugs in those patients.^(31, 35, 36)^ Deritis ratio (ALT/AST) typically ranges between 0.5-0.7 generally below 1.0 ^(37)^ In our results it was mostly higher side in almost all the patients with COVID-19 infection. We found a significant association between high Deritis ratio and disease mortality (P=0.018) that was also reported in some previous studies.^(36)^

In addition to LFTs the levels of CRP that is the marker of tissue damage and inflammation (acute phase protein) respectively^(38, 39)^ were found significantly high the patients with severe disease (CRP P=0.030) in our results. Zinellu et al also reported elevated levels of CRP (P<0.05) in severe patients ^(23, 32, 33, 36)^ So, these parameters can be used as predictive factors for developing cytokine storm/ severe disease and mortality risk.^(31)^

### Limitations

This study was single centered with a small number of patients so, cannot be generalized over the entire population. History of contact, past and current infections, treatment, intubation, and length of hospital stay, must be included to have accurate and complete data on all the risk factors associated with disease and progression towards severe disease. The lack of universal SARS-CoV-2 testing makes it difficult to identify whether people are truly asymptomatic or uninfected.

## Conclusion

Real-time PCR results for SARS-CoV-2 reported qualitatively (negative or positive) is sufficient for diagnosis but reporting of Ct values may have benefit for clinicians in patient management decisions. Still, more research is required to support this. Several risk factors for developing severe disease and mortality risk have been identified such as old age, underlying comorbidities such as diabetes and hypertension, etc. The relation between asthma and SARS-CoV-2 severity is not clear and needs further investigation. Deranged LFTs during infection are common and transient. Though, these laboratory parameters ALT, AST, Deritis ratio (AST/ALT), and CRP can be used as predictive biomarkers for progression towards severe disease and risk of mortality

### Suggestions

RT-PCR Ct value link with disease severity needs further investigation. So that, it can be used as a guideline for clinicians. As the data was taken only once, changes over the entire course of disease need to be closely monitored, with treatment history, to explore a more accurate relation between abnormal LFTs and the severity of disease. Further research is needed that either changes in liver enzymes are due to infection or due to treatment. Altogether, the data must be pooled and gathered across countries, so that we can optimize our understanding about the disease.

## Data Availability

erived data supporting the findings of this study are available from the corresponding author on request

## ACKNOWLEDGMENTS

I am grateful to Miss Saba Aziz for helping me in statistical analysis of data.

## Notes

### Competing Interest Statement

The authors have declared no competing interest.

### Funding Statement

This research received no specific grant from any funding agency in the public, commercial, or not-for-profit sectors.

